# Bacterial Profile and Antimicrobial Susceptibility Patterns of Asymptomatic Urinary Tract Infection Among Pregnant Women Attending Ante-natal Care at Kairuki Hospital, Dar es-Salaam, Tanzania

**DOI:** 10.64898/2025.12.18.25342602

**Authors:** Deogratias R. Gabriel, Ashura Mayanda, Anastasia Rogers, Hamis Kabuga, Seleman Kungulilo, Maureen Ngassala, Walter Msangi, Isaac Makanda, Monica Chiduo, Manento Mtango, Jeanne Pierre, Haji E Kapesa, Lemmygius D.Balilemwa, Magesa Mafuru

**Affiliations:** Department of Microbiology and Parasitology, Kairuki University, Tanzania; Department of Pharmacology and Therapeutics, Kairuki University, Tanzania; Department of Obstetrics and Gyanaecology, Kairuki Hospital, Tanzania; Microbiology Laboratory, Kairuki Hospital, Tanzania

**Keywords:** Pregnant women, Urinary tract infection, antimicrobial resistance, susceptibility testing

## Abstract

**Introduction:** Urinary tract infections are common to pregnant and nonpregnant women estimated to 150 million new cases annually. The incidence increases with pregnancy due changes that take place. Causative microbes are *E.coli, Klebsiella pneumoniae and Staphylococci species*. The disease presents symptomatically or asymptomatically, early investigation, detection and treatment to pregnant mothers are crucial to avoid maternal and foetal complications. Several effective antimicrobials are contraindicated using ineffective agents jeopardizes treatment outcome leading to multidrug resistance. We assessed UTI causative microbes and susceptibility patterns to common antibiotics.

**Methods:** We conducted a hospital based cross sectional study at Kairuki hospital involving 262 pregnant mothers attending ante-natal clinics. Mid-stream urine was collected and inoculated on Cysteine-lactose-electrolyte-deficient agar, MacConkey and blood agar. Eleven microbes were isolated and tested for susceptibility against antibiotics using Kirby-Bauer disk diffusion technique on Mueller-Hinton agar. Data analysed using SPSS package version 23.

**Results:** The prevalence of UTI in pregnant mothers was 31.2% (82/262). The gram positive isolates were more prevalent than gram nmoste (59.3% versus 40.7%) *Staphylococcus aureus* 22/82 (26.2%) and *S. saprophyticus* 15/82 (17.9%) were the mostly isolated. Nitrofurantoin, Piperacillin/tazobactam have lowest resistant rate to both gram negative and gram positive isolates ranging from (0-26%) while Erythromycin and Ampicillin have the highest resistant rate ranging from (60-100%) therefore associated with multidrug resistant.

**Conclusion:** Asymptomatic UTI is prevalent to pregnant women at this hospital. We recommend culture and sensitivity results to guide treatment and usage of nitrofurantoin, piperacillin/tazobactam as first line treatment of UTI in pregnancy.

## Background

Urinary tract infections (UTIs) are among common ailments in pregnant and nonpregnant women and the second pathological finding after anaemia. The prevalence of UTI range from 2-10% in different parts of the world, notably higher in Africa-American origin and lowest in whites with lower parity (1).The chance of developing UTI is four times higher in pregnant women than nonpregnant whilst the incidences are higher in developing world like Africa and Asia (2). Its estimated that the incidence approaches 150 million new cases per year (3). The UTIs are associated with anatomical, immunological and physiological changes which take place during pregnancy leading to urinary stasis and uretero-vesical reflux (4). Basically UTIs are caused by invading pathogens in the urinary tract leading to inflammation by ascending on the bladder, ureters and kidneys and other adjacent organs (5,6). If not promptly diagnosed and treated UTIs can lead to admission and likely to complicate to serious gynaecological and medical consequences such as abortions, intrauterine growth retardation, still births, preterm deliveries, maternal hypertension, pyonephrosis due to total kidney infection that can lead to capsule rupture hence development of perinephrosis and abscess to pregnant mothers and their babies (7). Clinically the diseases can present as either asymptomatic bacteriuria or symptomatic with acute cystitis, emphysematous pyelonephritis, perinephric abscess or acute kidney injury (8,9).

Generally the disease causative organisms are gastro-intestinal normal flora found in both pregnant and nonpregnant women, the commonest being the gram negative pathogens such as *E.coli* (80-90%), *Proteus mirabilis*, and *Klebsiella pneumoniae* while the gram positive such as *Staphylococci* and *Streptococci* species are less common (8,9). Empirical management protocols which are not guided by laboratory results are commonly exercised in many health settings. The younger maternal age and poor adherence to treatment guidelines by the patients are all key factors responsible for disease development and recurrence (12,13). In managing such situation the World Health Organization (WHO) recommends treatment of the UTI patients on the basis of culture and sensitivity testing (Antimicrobial Susceptibility testing) results, the same idea is advocated by the Tanzania government treatment guidelines (14) (STG). Therefore, failure in adherence to recommendations increases the risk of creating resistant microbes to commonly prescribed antimicrobials. Therefore, the aim of this study was to determine the prevalence of UTIs and assess their antimicrobial susceptibility patterns to commonly prescribed antibiotics. The findings of this study also advocate rational antibiotic use hence limiting emergence and spread of antibiotic resistance.

## Methodology

### Study design and Setting

This was a hospital based cross sectional study conducted from 1^st^ March to 31^st^ August 2023 at Kairuki hospital, a private tertiary and teaching hospital for Kairuki University (KU) and Kairuki School of Nursing (KSN) in Kinondoni municipal, Dar es salaam Tanzania. The hospital has a capacity of 150 beds, offering services to both inpatient and outpatients, preventive services for non-communicable diseases, and ante natal clinics where reproductive and child health (RCH) services are offered. Also, the hospital is a home for offering various specialities such as internal medicine, women’s reproductive health, paedriatrics and surgery.

### Study participants and sample size estimation

The study involved routine pregnant women attending at Kairuki hospital ante natal clinics from. All consented pregnant women who met inclusion criteria were enrolled in the study. But those who declined or had taken antibiotics within the past two weeks, had a recent history of medical instrumentation, or had immunocompromising conditions such as diabetes or HIV were excluded from the study. The minimum sample size for this study was 275, calculated using a formula for a single population proportion based on the prevalence of UTIs in Dar es salaam of 23% (15)error=5

### Ethical Clearance

An approval to carry out this study was granted by Kairuki University’s Institutional Research Ethics Committee (IREC) through letter Ref: No HKMU/IREC/27.10/165. The data collection permission was obtained from Kairuki hospital authority and oral informed consents were obtained from pregnant women attending ante natal clinics anonymity was maintained by using unique numbers to each participant instead of names.

### Data collection

We used pretested questionnaires to gather information on sociodemographic information, clinical details and UTI related symptoms.

### Sample collection and processing

During data collection exercise, doctors at clinics conducted face to face education to each participant on importance of the study, assurance of confidentiality, proper ways of sample collection, evaluation on the health status in relation to UTI from each participant and voluntary participation was strongly encouraged and the findings were recorded on the uniquely coded questionnaires so no names were used except unique numbers in maintenance of participants anonymity. Then after, participants were provided with sterile sample containers each uniquely labelled containing 0.5mg boric acid crystals (16),marked with date and time of sample collection then requested to fill in 10mils of mid-stream urine (MSU) after having cleansed genital areas with clean water and voiding initial urine. Then samples were brought at designated collection point for documentation. Eventually collected urine samples were taken to the Kairuki hospital laboratory for processing within one hour post collection. In case there was a delay in processing within that time the samples were refrigerated at 4°C to avoid multiplication of microbes in room temperature.

### Bacterial isolation and identification Culture and Sensitivity test

By using sterile wire loop urine specimens collected from each participant were inoculated on Cystein-Lactose-Electrocyte Deficient agar (CLED) (HiMEDIA^®^Maharashtra,India) Blood agar and MacConkey agar (HiMEDIA^®^Maharashtra,India) followed by streaking to allow discrete colonies of bacteria based on the standard microbiological procedures(17).

Inoculated agar plates were then aerobically incubated at 37 °C overnight. After 24 hours, the plates were examined for the growth of significant bacteria. Bacteriuria was confirmed based on the presence of >10^4^ colony-forming units per milliliter (CFU/mL), growth of one or two distinct microorganisms from the urine sample, and microscopic detection of 3–5 pus cells per high-power field, as described by Zboromyrska et al (18). In case of mixed growth a sterile subcultures were performed to get pure growth according to previous study (19).

Furthermore, the positive urine cultures were tested for their physical features such as colonies morphology, presence of hemolysis on blood agar and biochemical reactions. On addition to biochemical identification, we performed gram staining whereby Gram-positive bacteria had to undergo catalase test on pure colonies from which catalase positive were confirmed for *Staphylococci* and negative for *Streptococci*. We also differentiated between *Staphyloccocus aureus* and other *Staphyloccoci* species by performing coagulase test (16). In contrast gram negative bacteria were identified based on growing characteristics on (MacConkey media HiMEDIA^®^ Maharashtra,India) whereby bacteria were classified as lactose fermenters and non-lactose fermenters. Confirmatory of gram negative bacteria were also done by using API 20E identification profile index software (20).

### Antimicrobial Susceptibility Tests

Following identification, isolated microbes were subjected to antimicrobial susceptibility testing (AST) to the selected antibiotics commonly prescribed to treat UTI in pregnant patients (21). The AST was determined by using disc diffusion technique on Mueller-Hinton agar (MHA)( HiMEDIA^®^ Maharashtra, India) as per Kirby-Bauer disc agar diffusion technique (22), then results were obtained by measuring the diameter of inhibition zone and interpreted according to the Clinical laboratory Standards Institute guideline (23).

The measured diameters of bacterial growth inhibition zones around the disks to the nearest millimeter and the results were classified as susceptible (S), intermediate (I) or resistant (R).

The susceptibility tests were performed on the following eleven antibiotic disks(Liofilchem® s.r.l. Roseto degli Abruzzi, Italy) which are commonly prescribed at this hospital: Erythromycin (15μg), Ampicillin (10μg), Nitrofurantoin (300μg), Piperacillin/Tazabactam (110μg), Trimethoprime/Sulfamethoxazone (25μg), Ceftazidime (30μg), Penicillin-G (10IU), Cefotaxime (30μg), Gentamycin (30μg), Amoxycillin/Clavulacacid (30μg) and Cefepime (30μg).

### Data Management and Analysis

All necessary information on each questionnaire and laboratory examination results were carefully recorded on excel spread sheet and cleaned for typing errors. The demographics and baseline clinical characteristics were summarized in frequency distribution tables and bar charts. The prevalence of UTI and the proportional of susceptible uropathogen bacteria against the selected antibiotics were analyzed using descriptive statistics. Data were analyzed using SPSS package version 23.

## Results

### 1.1 Demographics and clinical characteristics of study participants

A total of 262 pregnant women with mean age of 31.7 ± 4.3 years attended antenatal clinic at Karuki hospital from March 2023 to August 2023 during study period were enrolled and provided urine samples. Many participants were in third trimester of gestation (44.7%) compared to second and first trimester, 36.3% and 19.1% respectively. About 85% of the women were married and had adequate knowledge on UTI. Miscarriage history was uncommon, and majorities were either self-employed / business or employed, with large number of members attained a college education level (73.3 %) (Data summarized on Table 1).

**Table 1.**
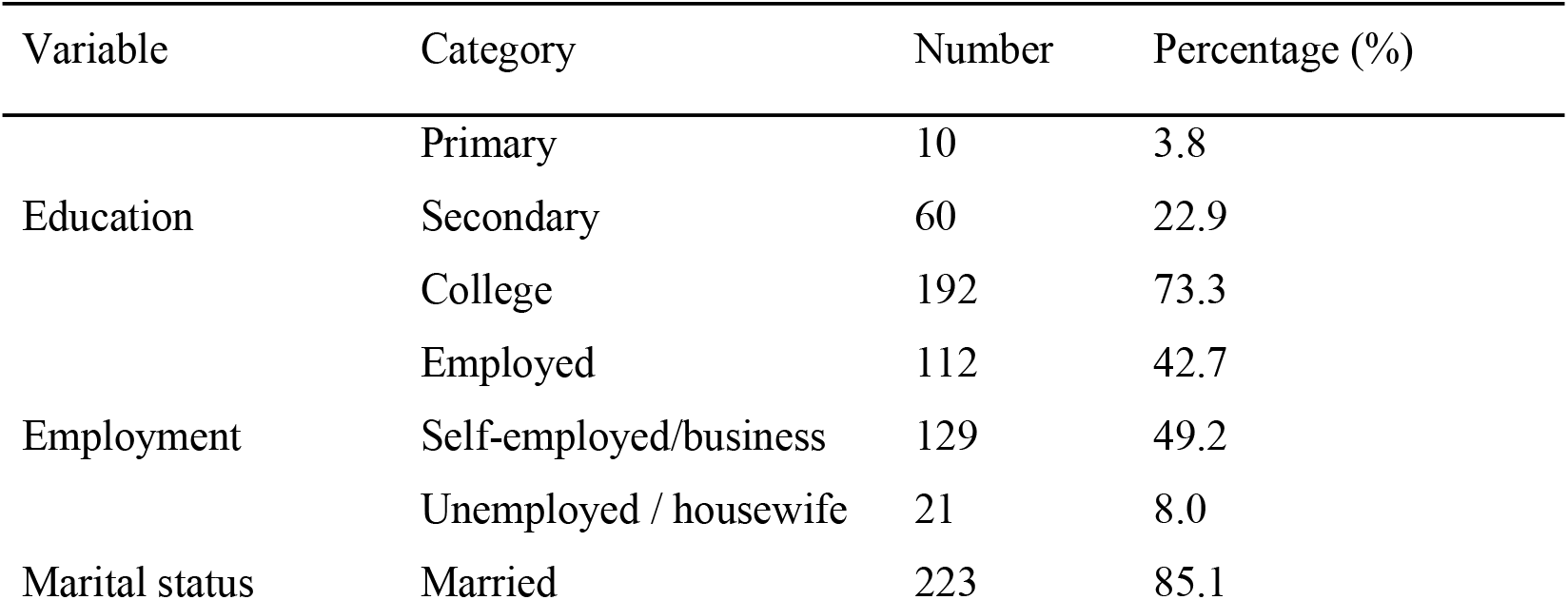
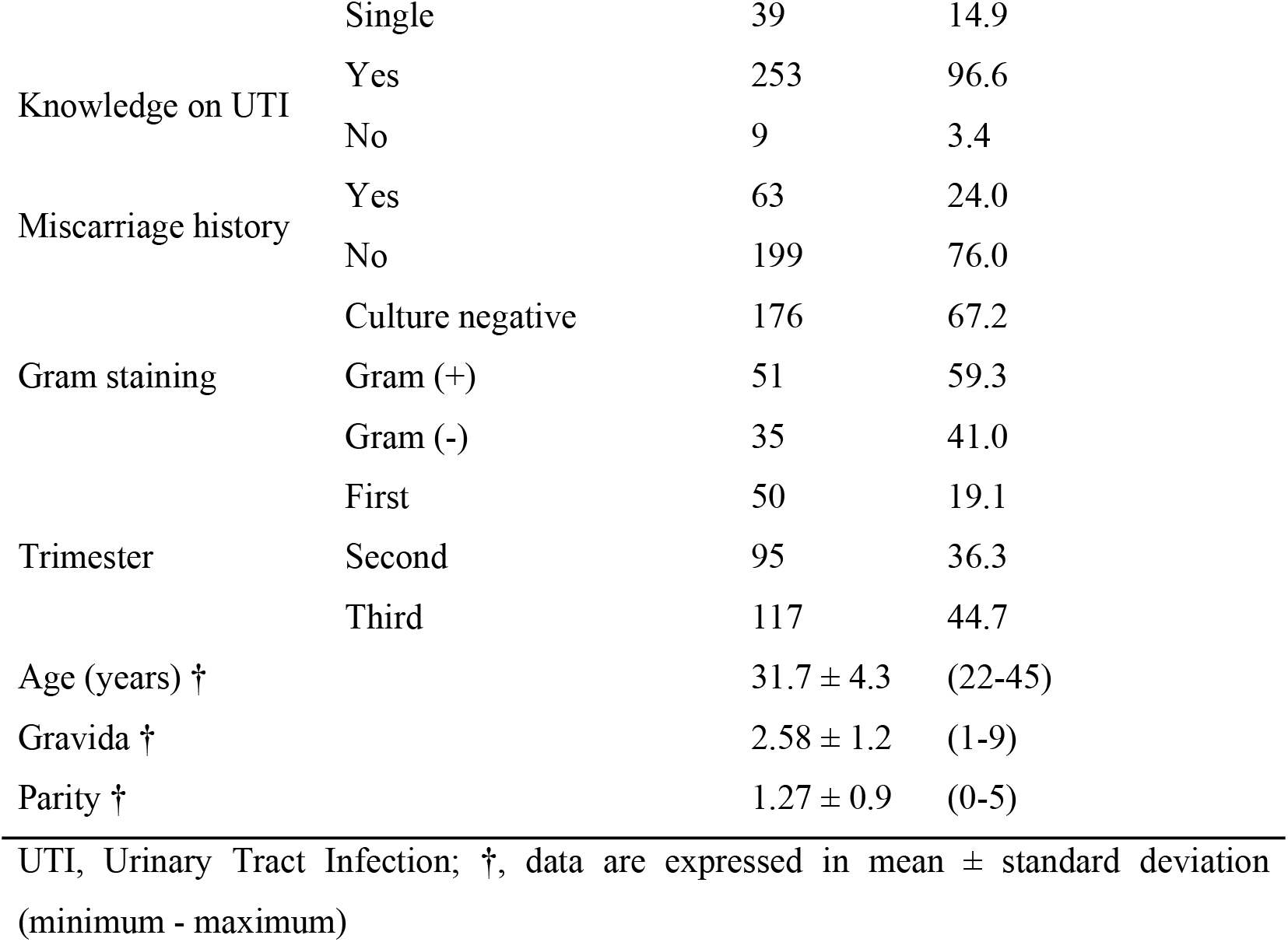
Demographics and clinical characteristics of study participants (n = 262)

### 1.2 The proportional of culture positive samples for different bacterial isolates

A mid-stream urine samples were collected and cultured for identification of different pathogenic bacterial isolates. Of 262 samples, 86 (33%) samples were culture positive (Figure 1). Among the culture positive samples, 59.3% of isolates were Gram positive while Gram negative comprised 41% of the isolates.

**Figure 1.**
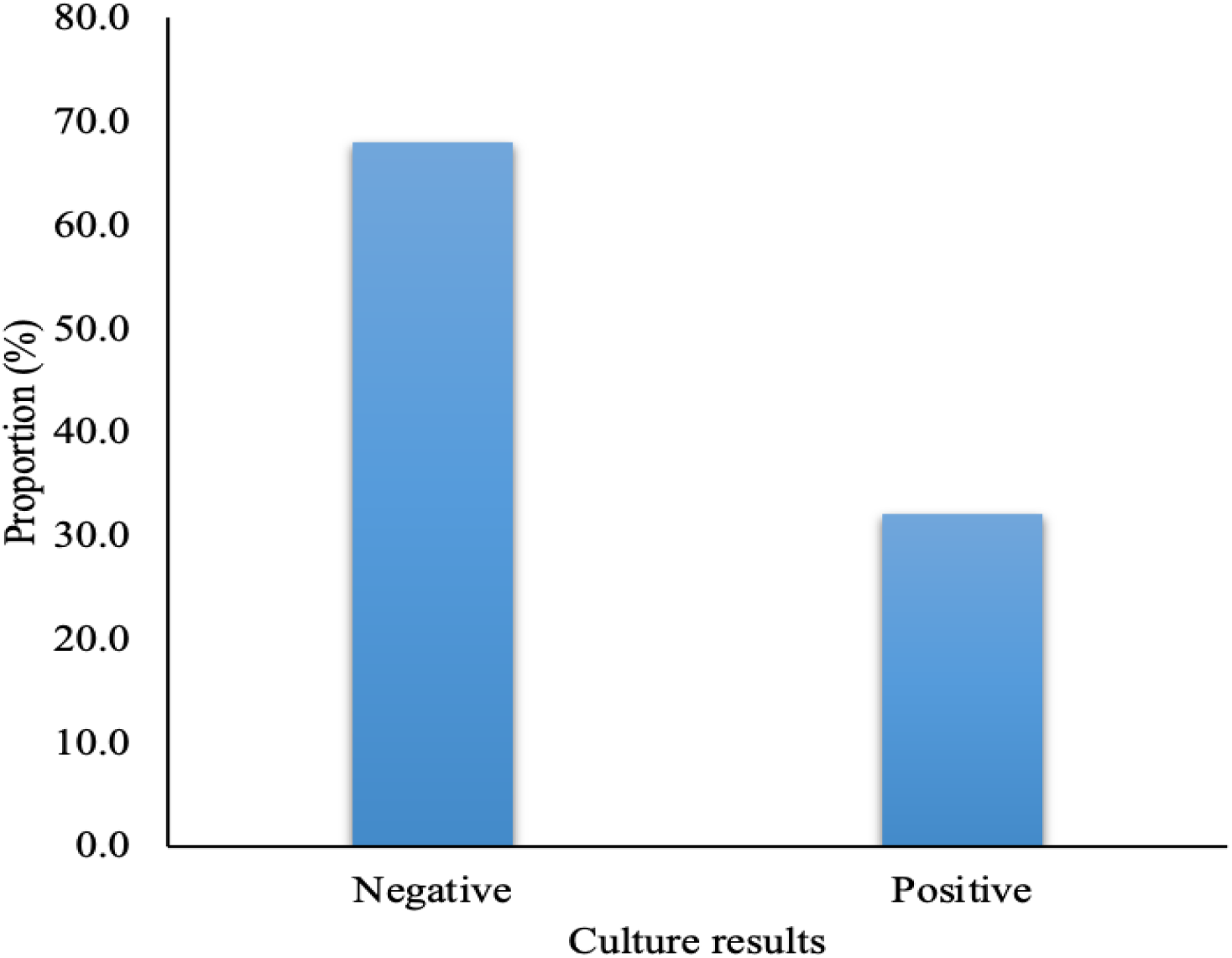
Culture results of the isolated samples (n = 262).

### 1.3 Bacterial isolates from the samples

Among 86 culture positive samples, a total of eleven (11) species of pathogenic bacteria were isolated of which gram positive were most common 51/86 (59.3%). The *Staphylococcus aureus* was the predominant isolate (25.6%) followed by *S. saprophyticus* (17.4%) The least isolated bacteria were gram negative *Enterobacter spp* and *Citrobacter freundii* at a proportion of 2.3% and 1.2% respectively (Table 2)

**Table 2.** Proportion of bacteria isolated from the samples (n = 86)

| Pathogens isolated | Samples | Percentage |
| --- | --- | --- |
| *Staphylococcus aureus* | 22 | 25.6 |
| *Enterococcus faecalis* | 5 | 6.0 |
| *Klebsiella. pneumoniae* | 13 | 15.5 |
| *Escherichia coli* | 8 | 9.5 |
| *Proteus spp* | 5 | 6.0 |
| *Staphylococcus. epidermidis* | 7 | 8.3 |
| *Ectobacteria spp* | 2 | 2.3 |
| *Serratia spp* | 3 | 3.6 |
| *Streptococcus agalactiae* | 3 | 3.6 |
| *Staphylococcus. saprophyticus* | 15 | 17.4 |
| *Citrobacter freundii* | 1 | 1.2 |

### 1.4 Antibiotic susceptibility pattern of isolated bacteria against selected antibiotics

The isolated bacteria were tested for their susceptibility to the commonly prescribed antibiotics using the Kirby-Bauer disc agar diffusion technique. The tested antibiotics were erythromycin, ampicillin, nitrofurantoin, piperacillin / tazobactam, sulfamethoxazole / trimethoprim, ceftazidime, penicillin-G, cefotaxime, gentamycin, amoxicillin / clavulanic acid and cefepime (Table 3 & Table 4).

**Table 3.**
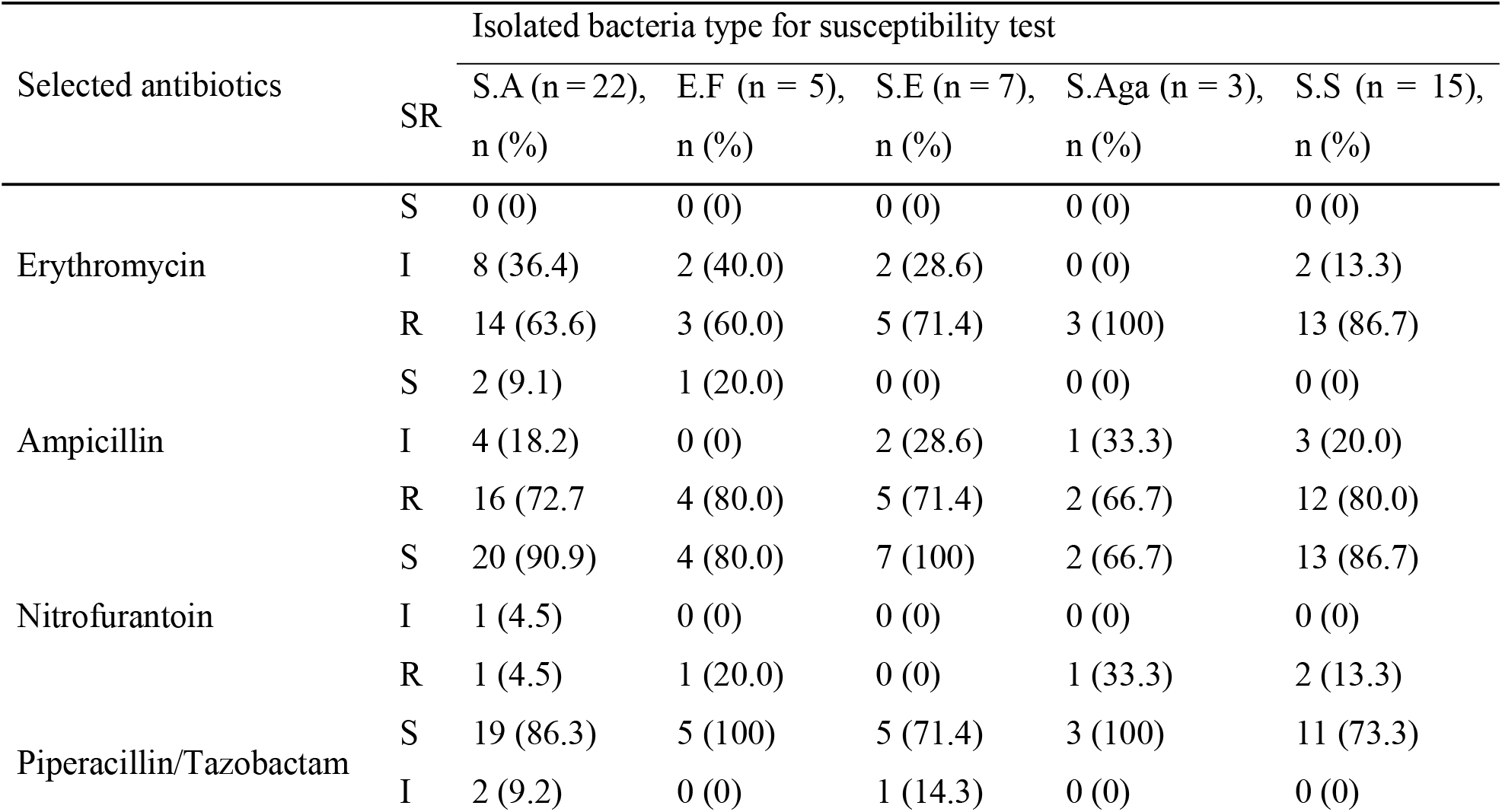
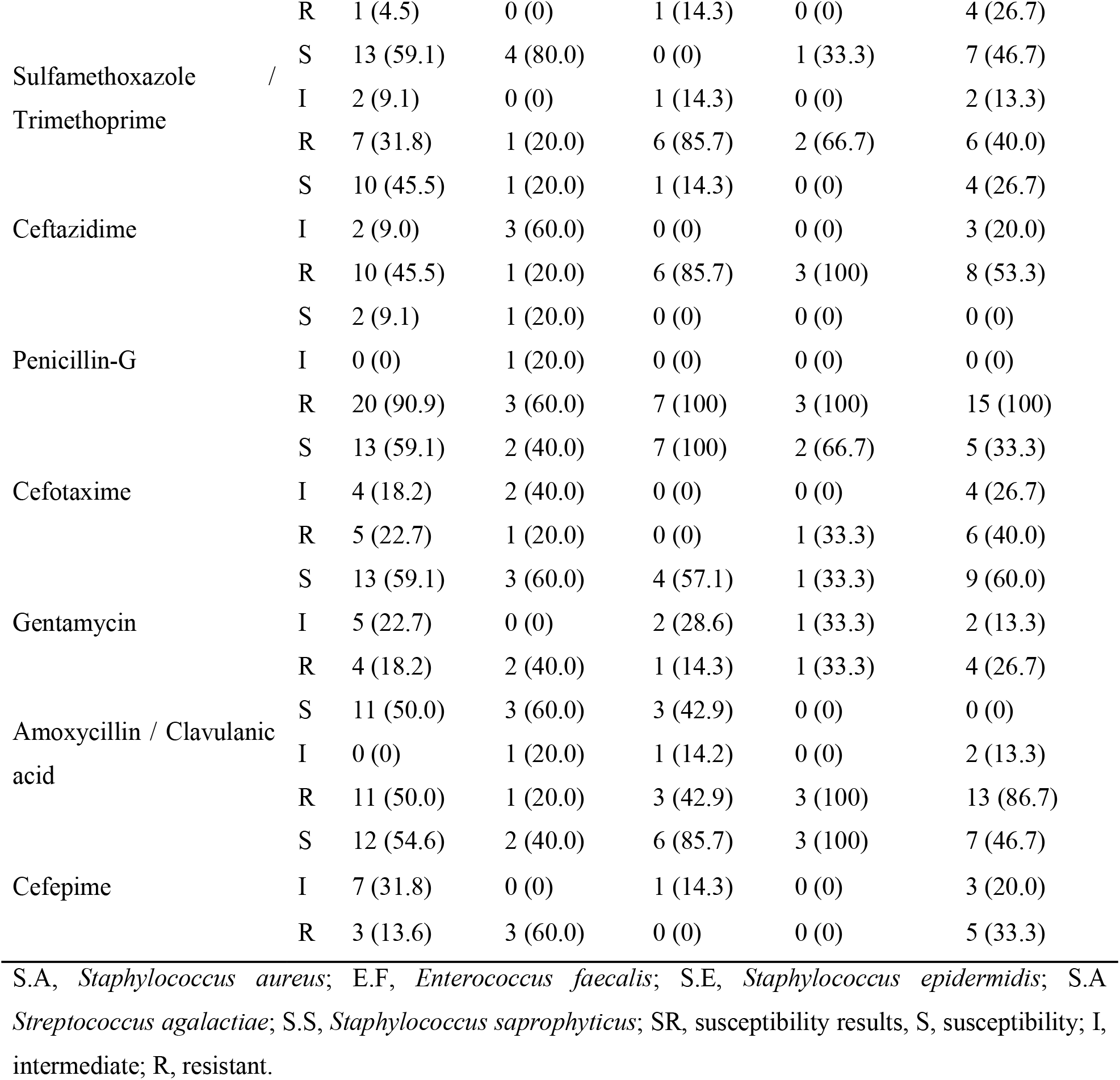
Antibiotic susceptibility pattern of isolated Gram-positive bacteria against selected antibiotics.

**Table 4.**
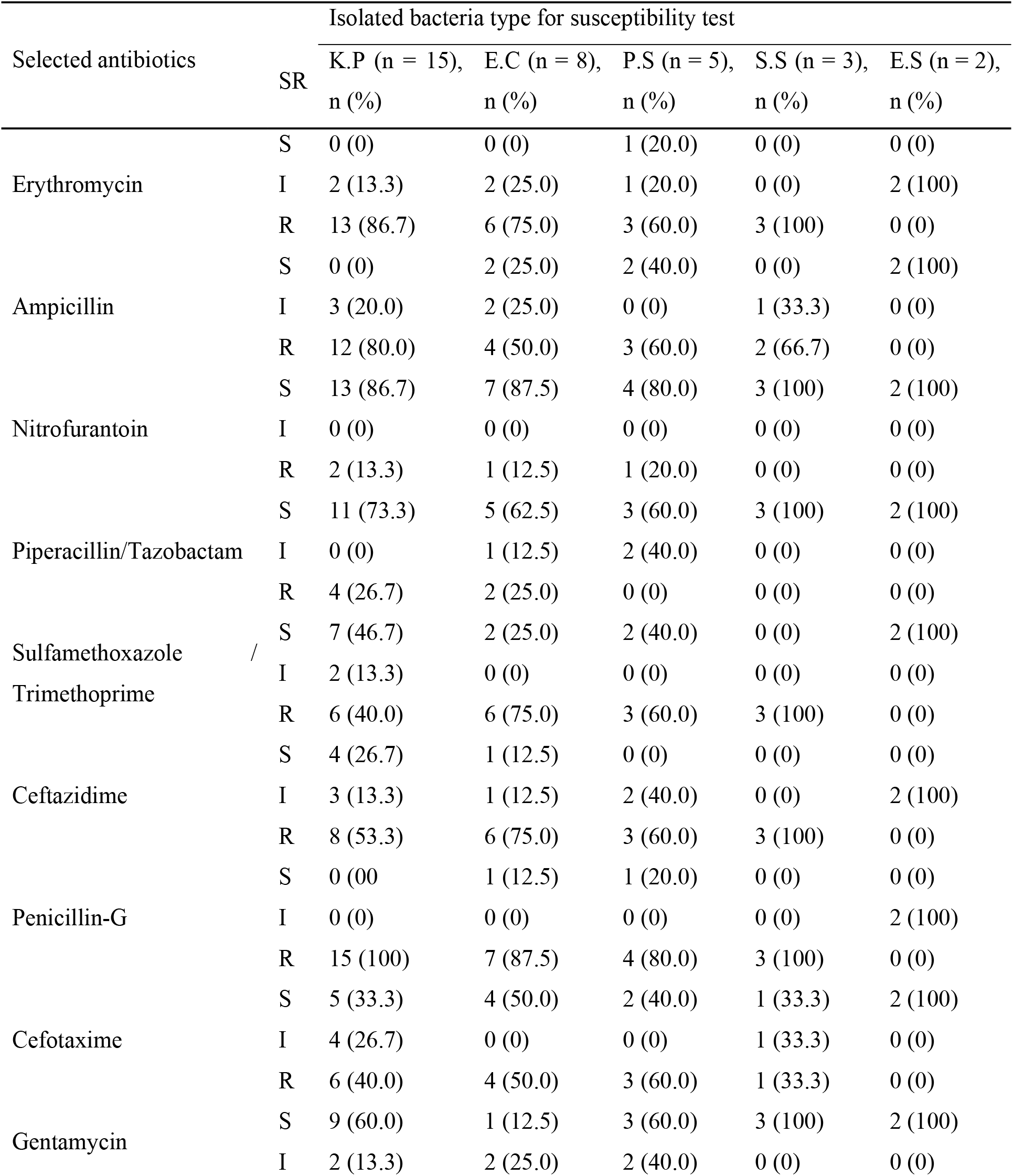
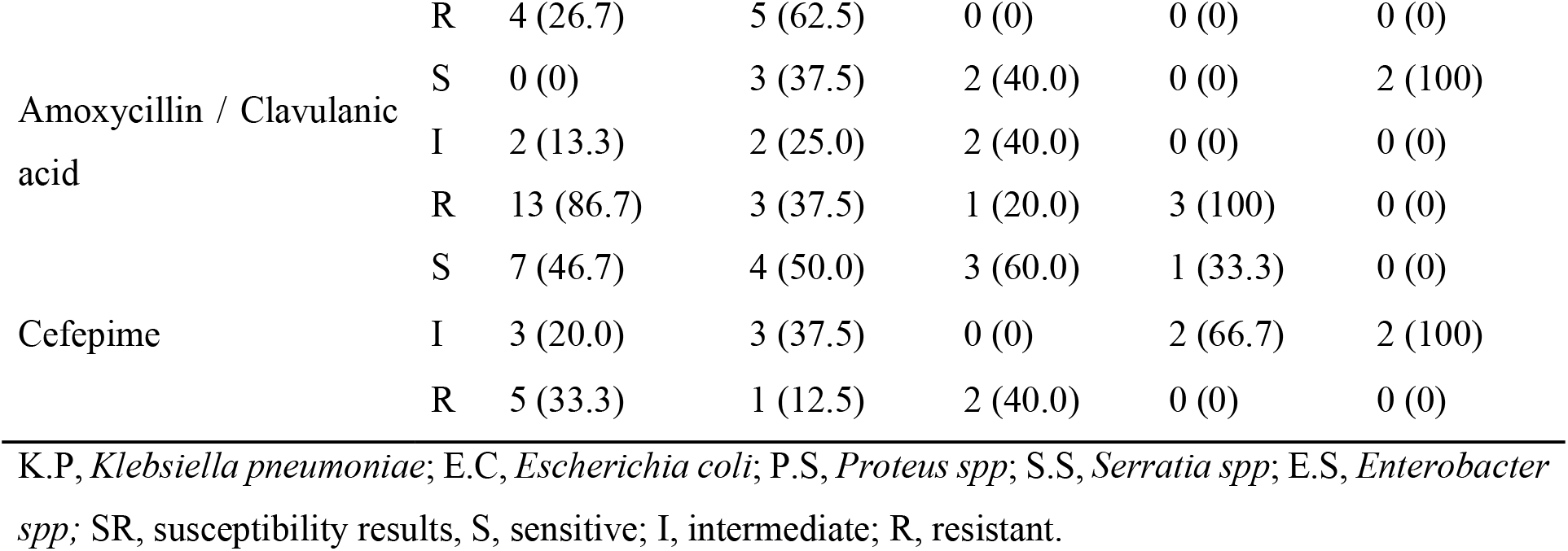
Antibiotic susceptibility tests of isolated Gram-negative bacteria on selected antibiotics.

## Discussion

This was the first study at this hospital to examine bacterial isolates profile and antimicrobial resistance patterns of urinary tract infection (UTIs) among pregnant women attending ante natal clinics, which revealed an overall prevalence of UTIs of 33% being two folds higher than the previous study done in Mwanza, Tanzania (17.9%) (24), Somaliland (16.4%)(25),,Eastern Ethiopia(14%) (26) Nairobi, Kenya14% (27) but lower than the study done in Morogoro, Tanzania (41%) (28), Central Uganda (73%) (29) and Ebonyi state, Nigeria(55%) (30). However the prevalence of UTIs in our study was generally similar to studies done in Mbarara regional referral hospital, South-Western Uganda (35%) (31) Ghana (39.8%) (32) and Tripoli Libya (37.3%)(33). In this study one could expect the lower prevalence of the infection due to education and economical status background of women who participated in this study but it was not the case, this could be due to the gestation age as the higher percentage of women enrolled in this study were in the 3rd trimester (34),but also the variation in prevalence can be the difference in sample size, social affiliations, personal hygiene habits and difference in geographical aspects.

Basically we didn’t notice any association of UTI and demographic factors like parity,age, education background and economical status of the subjects so our study findings correspond to the study done in Ethiopia (35). Among isolated bacteria, gram positive were more prevalent (59.3%)vs 40.7%) than gram negative. Corresponds to the study done in Ethiopia whereby gram positive were more common and *Staphylococcus aureus* predominated (22,26.2%)(36) followed by *S. saprophyticus*(15,17.9%) and gram negative *Citrobacter freundii* being the least (1,1.2%).In our findings Nitrofurantoin and Piperacillin/Tazobactam were sensitive to all gram positive bacteria so using them in empirical treatment especially when treating UTI in pregnancy is advisable. Similarly, Nitrofurantoin and Piperacillin/Tazobactam were also sensitive in gram negative bacteria while Gentamycin was more sensitive to *Serratia, Ectobacter* species, *Proteus, Klebsiella* but less sensitive to *E.coli*. Sulfamethoxazole / Trimethoprime and Cefotaxime were sensitive to *Ectobacter* species only so it should be avoided in treatment of diseases caused by the rest of bacterial species isolates this justifies treatment depending on culture and sensitivity results.

Erythromycin, Ampicillin and Penicillin G were not sensitive to any isolated gram positive bacteria whereas Sulfamethoxazole/Trimethoprime, Ceftaxidime and Amoxycillin/Clavulanic acid were insensitive to three isolated bacteria (MDR) therefore they pose ineffective outcome in management of UTI so should be avoided. However, Cefepime was insensitive to *E.faecalis* only so can be used in treating UTI patients caused by other bacteria though culture and sensitivity results remains to be important for appropriate management of UTI.

Erythomycin, Ampicillin, Salfamethoxazole/Trimethoprime, Ceftazidime, Pen-G and Amoxycillin were resisted by more than one gram negative bacteria, so they are no longer potent in treatment of UTI. Gentamycin and Cefotaxime are insensitive to only one isolate so can be used depending on culture and sensitivity results or complemented with other antibiotics which are sensitive.

## Conclusion

Based on the evidence from this study, UTI prevalence is high among pregnant women in 3^rd^ trimester, The gram positive isolates were more prevalent of which *S.aureus* seemed to be more predominant. Nitrofurantoin and Piperacillin/Tazobactam were the most sensitive antibiotics to both gram positive and gram negative isolates whereas other antibiotics such as Erythomycin, Ampicillin, Salfamethoxazole/Trimethoprime, Ceftazidime, Pen G and Amoxycillin were less or total insensitive to many isolates. Multi-drug resistance was appreciated to many isolates justifying incorporation of culture and susceptibility tests results to guide our routine management of UTI to pregnant mothers in avoiding of recurrence and creation of resistant microbes.

## Data Availability

All relevant data are within the manuscript and its Supporting Information files.

## Author contributions

**Conceptualization:** Deogratias R Gabriel, Hamis Kabuga

**Formal data analysis: Magesa** Mafuru, Deogratias R Gabriel, Ashura Mayanda

**Investigation and sample processing:** Hamis Kabuga,Maureen Ngassala, Seleman Kungulilo,Walter Msangi,Lemmygius Balilemwa,Haji E.Kapesa.

**Methodology and Data collection:** Isaac Makanda, Manento Mtango,Monica Chiduo,Jeanne Pierre **Njoli**

**Project administration:** Deogratias R Gabriel, Magesa Mafuru **Resources:** Ashura Mayanda, Annastasia Rodgers, Hamis Kabuga **Supervision:** Deogratias R Gabriel, Ashura Mayanda, Magesa Mafuru

**Original Manuscript Writing:** Deogratias R Gabriel, Ashura Mayanda, Magesa Mafuru

**Writing**,**review & editing:** Deogratias R Gabriel, Magesa Mafuru, Hamis Kabuga, Ashura Mayanda, Lemmygius Balilemwa, Annastasia Rodgers, Monica Chiduo, Maureen Ngassala

Critically all authors revised the manuscript and collectively agreed to be responsible for all aspects of this paper and for its publication

## Acknowledgments

Kairuki University partially supported this study by providing funds to purchase Api20E kit we real appreciate for the support. The authors would like to thank participants for their commendable participation in provision of information lastly we appreciate the contribution by Prof Titus Kabalimu for his constructive comments and advises on the manuscriptdevelopment also Mr Rayner Rithe contributed Zotero reference software installation and its application we appreciate for that.

## References

1. Onoh RC, Umeora OUJ, Egwuatu VE, Ezeonu PO, Onoh TJP. Antibiotic sensitivity pattern of uropathogens from pregnant women with urinary tract infection in Abakaliki, Nigeria. Infect Drug Resist. 2013;6:225–33.

2. Belete MA, Saravanan M. A systematic review on drug resistant urinary tract infection among pregnant women in developing countries in africa and asia; 2005-2016. Infect Drug Resist. 2020;13:1465–77.

3. Abou Heidar N, Degheili J, Yacoubian A, Khauli R. Management of urinary tract infection in women: A practical approach for everyday practice. Urol Ann. 2019;11(4):339–46.

4. Hannan TJ, Hooton TM, Hultgren SJ. Estrogen and Recurrent UTI: What Are the Facts? Sci Transl Med [Internet]. 2013 June 19 [cited 2025 Mar 17];5(190). Available from: https://www.science.org/doi/10.1126/scitranslmed.3006423

5. Belete MA, Saravanan M. <p>A Systematic Review on Drug Resistant Urinary Tract Infection Among Pregnant Women in Developing Countries in Africa and Asia; 2005–2016</p>. Infect Drug Resist. 2020 May;Volume 13:1465–77.

6. ajol-file-journals_54_articles_186034_submission_proof_186034-637-473011-1-10-20190429LEO.

7. Amiri M, Lavasani Z, Norouzirad R, Najibpour R, Mohamadpour M, Nikpoor AR, et al. Prevalence of Urinary Tract Infection Among Pregnant Women and its Complications in Their Newborns During the Birth in the Hospitals of Dezful City, Iran, 2012 - 2013. Iran Red Crescent Med J [Internet]. 2015 Aug 24 [cited 2025 May 30];17(8). Available from: https://archive.ircmj.com/article/17/8/16665-pdf.pdf

8. Kolman KB. Cystitis and Pyelonephritis. Prim Care Clin Off Pract. 2019 June;46(2):191–202.

9. Gilbert NM, O’brien VP, Hultgren S, Macones G, Lewis WG, Lewis AL. Urinary Tract Infection as a Preventable Cause of Pregnancy Complications: Opportunities, Challenges, and a Global Call to Action. Glob Adv Health Med. 2013 Sept;2(5):59–69.

10. Demilie T, Beyene G, Melaku S, Tsegaye W. URINARY BACTERIAL PROFILE AND ANTIBIOTIC SUSCEPTIBILITY PATTERN AMONG PREGNANT WOMEN IN NORTH WEST ETHIOPIA.

11. Kalinderi K, Delkos D, Kalinderis M, Athanasiadis A, Kalogiannidis I. Urinary tract infection during pregnancy: current concepts on a common multifaceted problem. J Obstet Gynaecol. 2018 May 19;38(4):448–53.

12. Barlow G, Nathwani D. Is antibiotic resistance a problem? A practical guide for hospital clinicians. Postgrad Med J. 2005 Nov 1;81(961):680–92.

13. Irunde H, Minzi O, Moshiro C. Assessment of Rational Medicines Prescribing in Healthcare Facilities in Four Regions of Tanzania. J Pharm Pract Community Med. 2017 Oct 15;3(4):225– 31.

14. United Republic of Tanzania. STANDARD TREATMENT GUIDELINES AND NATIONAL ESSENTIAL MEDICINES LIST FOR TANZANIA MAINLAND. United Republic of Tanzania; 2021.

15. Mwambete KD, Malaba P. High prevalence of antibiotic resistance among bacteria isolated from pregnant women with asymptomatic urinary tract infections in Dar es Salaam, Tanzania. Res J Health Sci. 2017 July 17;5(2):65.

16. Cheesbrough M. District laboratory practice in tropical countries: Part 2. Second edition. Cambridge: Cambridge University Press; 2006. 1 p.

17. M100-S11, Performance standards for antimicrobial susceptibility testing. Clin Microbiol Newsl. 2001 Mar;23(6):49.

18. Zboromyrska Y, Rubio E, Alejo I, Vergara A, Mons A, Campo I, et al. Development of a new protocol for rapid bacterial identification and susceptibility testing directly from urine samples. Clin Microbiol Infect. 2016 June;22(6):561.e1–561.e6.

19. Zboromyrska Y, Rubio E, Alejo I, Vergara A, Mons A, Campo I, et al. Development of a new protocol for rapid bacterial identification and susceptibility testing directly from urine samples. Clin Microbiol Infect. 2016;22(6):561.e1–561.e6.

20. Reynolds J. Contributors and Attributions.

21. Gajic I, Kabic J, Kekic D, Jovicevic M, Milenkovic M, Mitic Culafic D, et al. Antimicrobial Susceptibility Testing: A Comprehensive Review of Currently Used Methods. Antibiotics. 2022 Mar 23;11(4):427.

22. Kirby-Bauer-Disk-Diffusion-Susceptibility-Test-Protocol-pdf (1).

23. CLSI-2020.

24. Massinde A, Gumodoka B, Kilonzo A, Mshana S. Prevalence of urinary tract infection among pregnant women at Bugando Medical Centre, Mwanza, Tanzania. Tanzan J Health Res. 2009 July 1;11:154–9.

25. Ali AH, Reda DY, Ormago MD. Prevalence and antimicrobial susceptibility pattern of urinary tract infection among pregnant women attending Hargeisa Group Hospital, Hargeisa, Somaliland. Sci Rep. 2022 Jan 26;12(1):1419.

26. Tula A, Mikru A, Alemayehu T, Dobo B. Bacterial Profile and Antibiotic Susceptibility Pattern of Urinary Tract Infection among Pregnant Women Attending Antenatal Care at a Tertiary Care Hospital in Southern Ethiopia. Mastroeni P, editor. Can J Infect Dis Med Microbiol. 2020 Dec 24;2020:1–9.

27. Onyango HA, Ngugi C, Maina J, Kiiru J. Urinary Tract Infection among Pregnant Women at Pumwani Maternity Hospital, Nairobi, Kenya: Bacterial Etiologic Agents, Antimicrobial Susceptibility Profiles and Associated Risk Factors. Adv Microbiol. 2018;08(03):175–87.

28. Mlugu EM, Mohamedi JA, Sangeda RZ, Mwambete KD. Prevalence of urinary tract infection and antimicrobial resistance patterns of uropathogens with biofilm forming capacity among outpatients in morogoro, Tanzania: a cross-sectional study. BMC Infect Dis. 2023 Oct 5;23(1):660.

29. Enock K, Nakalema M. PREVALENCE OF URINARY TRACT INFECTION AMONG PREGNANT WOMEN ATTENDING ANTENATAL CLINIC AT KASANGATI HEALTH CENTER IV IN WAKISO DISTRICT. A DESCRIPTIVE CROSS-SECTIONAL STUDY. [Internet]. sjhr; 2023 [cited 2025 Apr 11]. Available from: https://sjhresearchafrica.org/index.php/public-html/article/view/451

30. Chukwudozie Onuoha S. Prevalence and Antimicrobial Susceptibility Pattern of Urinary Tract Infection(UTI) among Pregnant Women in Afikpo, Ebonyi State, Nigeria. Am J Life Sci. 2014;2(2):46.

31. Johnson B, Stephen BM, Joseph N, Asiphas O, Musa K, Taseera K. Prevalence and bacteriology of culture-positive urinary tract infection among pregnant women with suspected urinary tract infection at Mbarara regional referral hospital, South-Western Uganda. BMC Pregnancy Childbirth. 2021 Dec;21(1):159.

32. Vicar EK, Acquah SEK, Wallana W, Kuugbee ED, Osbutey EK, Aidoo A, et al. Urinary Tract Infection and Associated Factors among Pregnant Women Receiving Antenatal Care at a Primary Health Care Facility in the Northern Region of Ghana. Grande R, editor. Int J Microbiol. 2023 June 2;2023:1–10.

33. Hamida A, Amal A. Urinary Tract Infections among Pregnant Women at Tripoli. Int Arch Med Microbiol [Internet]. 2023 Dec 31 [cited 2025 Apr 14];5(1). Available from: https://clinmedjournals.org/articles/iamm/international-archives-of-medical-microbiology-iamm-5-019.php?jid=iamm

34. Cheung KL, Lafayette RA. Renal Physiology of Pregnancy. Adv Chronic Kidney Dis. 2013 May;20(3):209–14.

35. Alemu A, Moges F, Shiferaw Y, Tafess K, Kassu A, Anagaw B, et al. Bacterial profile and drug susceptibility pattern of urinary tract infection in pregnant women at University of Gondar Teaching Hospital, Northwest Ethiopia. BMC Res Notes. 2012 Dec;5(1):197.

36. Tadesse E, Teshome M, Merid Y, Kibret B, Shimelis T. Asymptomatic urinary tract infection among pregnant women attending the antenatal clinic of Hawassa Referral Hospital, Southern Ethiopia. BMC Res Notes. 2014;7(1):155.

